# Brain and vascular integrity related to cognitive and motor flexibility in autism: a study protocol

**DOI:** 10.64898/2026.06.24.26356429

**Authors:** Erik Domellöf, Andreas Johansson, Sara Stillesjö, Linnea Karlsson Wirebring, Carola Wiklund-Hörnqvist, Anna-Maria Johansson, Thomas Rudolfsson, Anders Wåhlin, Gustaf Wadenholt, Malin Ekesryd Nordström, Daniel Säfström

## Abstract

**Introduction:** Autism spectrum disorder, or autism, is a common neurodevelopmental condition characterized by socio-communicative problems together with restrictive and repetitive behaviors. Typically, the latter is manifested as deficits in behavioral flexibility, i.e. changing routine behaviors to adapt to environmental changes. Despite noticeable difficulties with flexible behavior in autism, there is to date not adequate knowledge about the intricacies of such challenges and neurobiological processes that may subserve them. This study aims to investigate both cognitive and motor flexibility in autistic compared with neurotypical adults using a novel combination of detailed methods for brain imaging and behavioral investigations in relation to probabilistic reversal learning (PRL) paradigms. In addition, the experiences of autistic adults on flexible behavior in education and everyday activities will be explored.

**Methods and analysis:** Differences in cognitive flexibility between autistic (n≥20) and neurotypical (n≥20) adults (18-35 years) will be investigated in terms of brain activations, measured by functional magnetic resonance imaging (fMRI), during two-choice PRL performance (cognitive task). In addition, group differences in microcirculation as measured by arterial spin labelling (ASL) will be evaluated. Group differences in motor flexibility will be investigated as expressed in movement planning and execution (spatio-temporal parameters), measured by a robotic manipulandum platform (KinArm End-Point Robot), during two-choice PRL performance (motor task). Semi-structured interviews will be conducted individually with autistic participants (n=15). Questions concern own experiences of cognitive and motor behavior, and strategies used to support flexibility in these behaviors. Data from this qualitative approach will be analyzed by thematic analysis.

**Ethics and dissemination:** Ethical approval has been obtained from the Swedish Ethical Review Authority (ref:2025-07939-01) and the study will be conducted in accordance with the Declaration of Helsinki, the European Union General Data Protection Regulation (GDPR) and national guidelines for the storing of personal data. The different investigations included are well-established, non-invasive and safe. Study outcomes will be published in peer-reviewed international scientific journals (open access), presented at national and international conferences, and to any interested audience/stakeholders.

**STRENGHTS AND LIMITATIONS OF THIS STUDY:**

- High-resolution measurements will be used to collect data on the nature of both cognitive and motor flexibility in autistic adults within a narrow age range.
- The study is expected to generate novel data revealing the links between brain activation, cerebral blood flow, and ability for flexible behavior in autism.
- Includes the voices of autistic adults themselves regarding experiences of flexible behavior in educational and leisure contexts.
- Flexible behavior in quantitative data collection is limited to performance on probabilistic reversal learning tasks.

## INTRODUCTION

With a global prevalence of ∼1% or higher,^1^ autism spectrum disorder, or autism, is one of the most common neurodevelopmental conditions in society. Autism is typified by social and communicative impediments and restricted behaviors, contributing to functional difficulties in everyday life.^2^ Among the significant challenges affecting individuals with autism are difficulties with sensory processing, motor performance, executive functioning, learning, anxiety, and unemployment.^1^

Consistent with the well-known restrictive and stereotypic behaviors, autism is also typified by deficits in behavioral flexibility.^3^ Although many everyday tasks are performed nearly automatically, it is sometimes necessary to change automatic behaviors if something changes in our environment and the behavior becomes inadequate. Thus, deficits in behavioral flexibility may contribute to difficulties adapting to novel situations and environments. For example, a predictable environment often provides security for autistic people, and a person with autism may function normally in a familiar environment but can decompensate in a challenging environment such as a grocery store. Impairments in behavioral flexibility might also be related with learning problems, since updating of behaviors typically is based on novel experiences.^4^ Given that the challenges affecting autistic individuals range from the cognitive to the motor domain, and that learning from novel experiences is crucial across domains, impairments in behavioral flexibility could be at the core of many difficulties displayed in everyday life for autistic individuals.

While it is recognized that autism originates from complex bidirectional gene-environment interactions in utero preceding an atypical development of neural networks,^1^ it is not yet fully understood how critical neurobiological processes may influence the unique challenges experienced by autistic individuals later in life. To date, several neuroimaging studies have demonstrated structural differences in the autistic brain, atypical brain activation patterns in relation to functional tasks, and atypical brain network functional connectivity.^5–8^ Thus, this literature has provided critical insight into neurobiological processes subserving various behavioral issues in autism at various ages. Still, less is known about the neural foundation of the prominent difficulties with behavioral flexibility in autism. There is evidence that both cognitive and motor flexibility are importantly subserved by fronto-striatal neural networks in the brain.^9–10^ The nature of this brain-behavior relation in autism is however not adequately investigated.

Reversal learning paradigms are a means to investigate aspects of cognitive flexibility by studying cognitive processes associated with reward-guided response selection and evaluation of responses depending on external feedback, well suited to use in neuroimaging studies.^11^ All reversal learning tasks involve the requirement to inhibit a prepotent but no longer appropriate response to a previously rewarded stimulus and a change of behavior towards a new response-reward likelihood (i.e. cognitive flexibility). Probabilistic reversal learning (PRL) is a more complex version of reversal learning where sometimes the correct response is not rewarded, i.e. further challenging the certainty of a response. Thus, successful performance in PRL tasks requires model-based strategies to cope with the probabilistic errors.

Neuroimaging studies involving various forms of reversal learning have been carried out in both healthy and clinical human populations (such as schizophrenia, depression, addiction, Parkinson’s disease, amyotrophic lateral sclerosis, dementia and patients with surgically excised brain lesions) and animals. Across species, the orbito-frontal cortex (OFC), and to some extent the medial prefrontal cortex (mPFC), have been identified as important cortical regions for reversal learning performance. In addition, at the subcortical level, dorsal and ventral regions of the striatum along with the amygdala are typically implicated.^11^ Thus, a neural circuitry involving the OFC, striatum and amygdala appears to be engaged during reversal learning. In addition, PRL seems to further engage the ventrolateral prefrontal cortex.^12^

Of relevance to the present project is that, although variants of reversal learning have been applied to study the neural basis of cognitive flexibility in different clinical populations, there are surprisingly few studies that have investigated autistic individuals. Thus, detailed investigation of the neurocognitive processes involved in performing reversals in autistic individuals has the potential to significantly advance the understanding of disrupted ability for rapid behavioral change in autism, of great relevance for clinical and educational interventions. Importantly, PRL tasks may help to further specify the impairments in behavioral flexibility in autism. For instance, both an insensitivity to feedback and an overly rigid internal model may contribute to reduced behavioral flexibility. Thus, PRL tasks during neuroimaging are well-suited for describing aspects of cognitive (in)flexibility and the underlying cortical and subcortical mechanisms.

According to a recent review,^13^ there is only a limited number of neuroimaging studies (N = 9) available measuring activation differences between autistic and neurotypical individuals during performance of any type of cognitive flexibility task. To date, only two neuroimaging studies have investigated this in autistic populations, both using PRL paradigms during functional magnetic imaging resonance (fMRI). In the first study,^14^ it was found that autistic male adolescents displayed reduced activity in mPFC and precuneus during PRL performance. The second study^15^ found reduced engagement in brain regions assisting cognitive decision-making processes (frontal motor planning systems, parietal cortex and the anterior cingulate cortex) in young autistic compared with neurotypical individuals. In addition, reduced activation was reported in brain regions of importance for reinforcement learning processes (ventral striatum and affective sector of the anterior cingulate cortex). The reduced mPFC activity in autistic youth during reversal learning involving probabilistic reinforcement found by Chantiluke and colleagues^14^ was also replicated.

An important, although often overlooked, aspect in autism is the cerebral vasculature subserving brain function. In relation to its comparatively small volume in terms of body mass, the brain requires a significant amount of body energy to function properly.^16^ Therefore, the vasculature of the brain plays a central supportive role for the neural cells and safeguards typical brain maturation, development and function across the lifespan.^16^ Since autism is a neurodevelopmental condition, the integrity of brain vasculature emerges as a particularly important factor to consider as a neurobiological underpinning in relation to challenges faced by autistic individuals.

Previous studies have shown deviations in the trajectories of brain development in children with autism, including measures of perfusion.^17^ Most perfusion studies report hypoperfusion (i.e. insufficient blood flow) in many brain regions in autistic compared with neurotypical individuals,^18^ although selective hyperperfusion also has been reported.^19–20^ Using single photon emission computed tomography (SPECT), Wilcox et al.^21^ found significant hypoperfusion in prefrontal areas in autistic individuals. In another SPECT-study, Ohnishi et al.^22^ found that perfusion abnormalities were related to specific cognitive dysfunctions, with correlations between autistic symptoms (such as impairments in social interaction) and abnormal patterns of perfusion in the limbic system and the mPFC. More recently, studies of cerebral blood flow (CBF) using non-invasive arterial spin labeling (ASL) imaging to detect changes in cerebral hemodynamics in autistic samples have emerged. Many of these studies focus on children and show widespread deviances in CBF in autism. Hypoperfusion is a common finding^23–27^ which appear to be non-linearly increasing with age, with a cross-sectional comparison showing fewer brain regions involved in toddlerhood than in adolescence.^25^ However, not all ASL studies in autism show regional or global hypoperfusion.^19, 25, 28^ Higher CBF has been shown in frontal white matter and subcortical gray matter in autism^20^ and one study^19^ reports both hyper- and hypoperfusion in regions of the medial frontoparietal network (default mode network). Regional hyper- and hypoperfusion have been associated with level of social difficulties^20^ and symptom severity^19^ in autism, and regional hypoperfusion have been linked to face recognition difficulties.^28^

Notably, most ASL studies include children and adolescents and only one study^20^ also involves adults. The age spans are typically large and some even include children as young as 2 years of age. Given the differences in developmental trajectories in CBF reported in autism^25^ and the typically wide age ranges, a focused study including only adults within a narrow age range might yield less ambiguous data as developmental aspects are limited. Us knowingly, there are no studies using ASL in autism that focus solely on adult samples with narrower age ranges. Such an enterprise may significantly further the knowledge in this inconclusive field of study.

The present multidisciplinary project aims to investigate behavioral flexibility in autistic adults compared with neurotypical adults of comparable age and sex. The project involves a novel combination of detailed methods for brain imaging and behavioral investigations of cognitive and motor flexibility using PRL paradigms. In addition, semi-structured interviews will be conducted with a sample of the participating autistic adults on their perspective of flexible behavior in education and everyday activities, including motor function. The results are expected to add importantly to current knowledge of adverse neurobiological processes and abnormalities in the vascular integrity of the brain in autism, including behavioral manifestations and individual experiences, and guide best clinical practice for daily life functioning.

### Study aims

Both cognitive and motor flexibility will be studied using PRL paradigms. Thus, while PRL mainly has been used in relation to cognitive performance, the present project will further introduce the paradigm in a motor task (i.e. motor flexibility).

In one session, brain activations will be measured by fMRI producing blood oxygen level dependent (BOLD) contrasts for activity in various brain regions during two-choice PRL performance (cognitive task). Main aims are to investigate group differences (autistic vs neurotypical) in contingency reversals and probabilistic errors as expressed in both behavioral and brain activity data.

In adjunct, the integrity of the microcirculation will be assessed with ASL. Differences between autistic and neurotypical adults will be investigated to characterize vascular integrity and to reveal any relation between perfusion and fMRI data.

In a separate session, outside of the MR-scanner, movement planning and execution will be measured by a robotic manipulandum platform (KinArm End-Point Robot) during two-choice PRL performance (motor task). The main aim of these experiments is to investigate differences between autistic and neurotypical adults in how contingent relationships between sensory stimuli and motor behavior (sensorimotor “rules”) are established, retained and updated. Reduced flexibility may result in difficulties with updating sensorimotor rules when the required relation between sensory stimuli and motor behavior changes.

A third session consists of semi-structured interviews to be conducted individually with autistic participants. The aim of this qualitative approach is to include the voices of autistic adults themselves regarding experiences both in school and in leisure contexts, in relation to flexible behavior, adding a dimension that is not possible to capture by quantitative methods. The respondents will be asked to describe both their current situation and how it has been previously in life. Interview questions cover experiences of cognitive and motor behavior, as well as own strategies participants use to support flexibility in these behaviors, for example when learning new motor skills and in situations where contextual conditions suddenly change.

Additional aims include investigating outcomes and group differences from behavioral measures collected by standardized neuropsychological and motor assessments (fluid intelligence, working memory, cognitive planning, set shifting, fine motor performance), also in relation to measures and task outcomes from the main investigations described above.

### Hypotheses

We hypothesize that, as compared with neurotypical individuals, individuals with autism will show deficits in cognitive and motor flexibility in the PRL tasks. We also hypothesize that these deficits will be accompanied by reduced activations and hypoperfusion in specific brain areas, where fronto-striatal neural networks may be particularly affected. Outcomes from the semi-structured interviews are expected to capture autistic individuals’ own perspective of flexible behavior in education and everyday activities. This will complement the project with a deeper understanding of how autistic adults themselves experience cognitive and motor flexibility, and cope with maneuvering in an unpredictable environment.

## METHODS AND ANALYSIS

### Study design

As outlined in the study aims, three steps for investigation are proposed. The first step, based on discussions with members of Umeå Center for Functional Brain Imaging (UFBI; https://www.umu.se/ufbi/), is a cross-sectional study of differences and similarities between autistic and neurotypical adults in brain activation patterns during a two-choice PRL paradigm (cognitive flexibility) by fMRI, together with structural CBF assessment by ASL. The second step involves a cross-sectional approach to investigate differences and similarities between autistic and neurotypical adults in spatio-temporal movement parameters during a two-choice PRL paradigm (motor flexibility) by KinArm. The third step is a qualitative approach involving semi-structured interviews of autistic adults. At present, participant recruitment and data collection are about to start. Thus, as requested for a study protocol and in compliance with BMJ Open policy, no data has been included. Fig 1 outlines the timeline for the project (Gantt chart).

**Figure 1.**
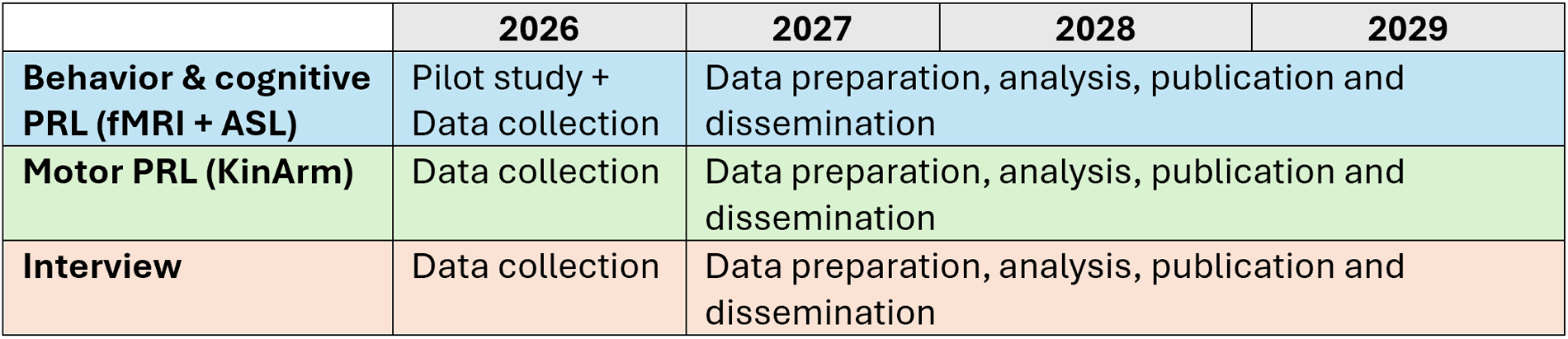
Project timeline

### Participants

A total of 20 autistic and 20 neurotypical adults (18-35 years) will be recruited for the fMRI (step 1) and KinArm (step 2) investigation, respectively. A total of 15 autistic adults (18-35 years) will be recruited for the interview (step 3). Numbers of participants are in keeping with previous similar studies using fMRI,^29^ the proposed motor task,^30–31^ and interview studies.^32^ Sample sizes can thus be considered acceptable to generate statistically significant outcomes (step 1-2) and for saturation in qualitative research (step 3). Inclusion criteria for the neurotypical adults are Swedish language fluency, normal or corrected-to-normal vision, and no known neuropsychiatric or motor disability. Inclusion criteria for the autistic adults are an autism spectrum disorder diagnosis established within the Swedish health care system, no diagnosed intellectual disability, normal or corrected-to-normal vision, and Swedish language fluency. For both groups, exclusion criteria for the fMRI study are non-compliance with the testing protocol, metal objects implanted (or otherwise) in the body, or pregnancy. For the KinArm study, non-compliance with the testing protocol will lead to exclusion. Participants are recruited through advertising in the Umeå region, by re-recruitment of participants from a previous similar study by the research group, and by convenience sampling. Written, informed consent is collected from all participants.

### Project description

#### 1. fMRI and CBF measurements

Brain activations supporting cognitive flexibility will be investigated by a 3 Tesla MR scanner at UFBI.

##### PRL task

In the first step, brain activation patterns during a computerized PRL-based cognitive task will be studied in the participants (autistic vs neurotypical). The task, an adaptation of previously used PRL paradigms,^14–15, 33^ will be performed in the MR scanner. Participants will see a computer screen via a mirror attached to the head coil, and responses will be collected through a four-button response pad. See Fig 2 for layout of the task design. On each trial, an image of two identical closed doors (one on the left, one on the right) is presented for 2 s on the screen and participants are asked to choose one by pressing the corresponding button within this time limit. The chosen door is then opened to reveal outcome feedback for 1.5 s, in the form of a green check mark indicating positive/reward/correct and a red cross indicating negative/non-reward/incorrect. After a semi-randomized inter-trial interval (ITI) of 0.5-7 s (mean = 2.5 s, weighted toward shorter interval), the next trial begins.

**Figure 2.**
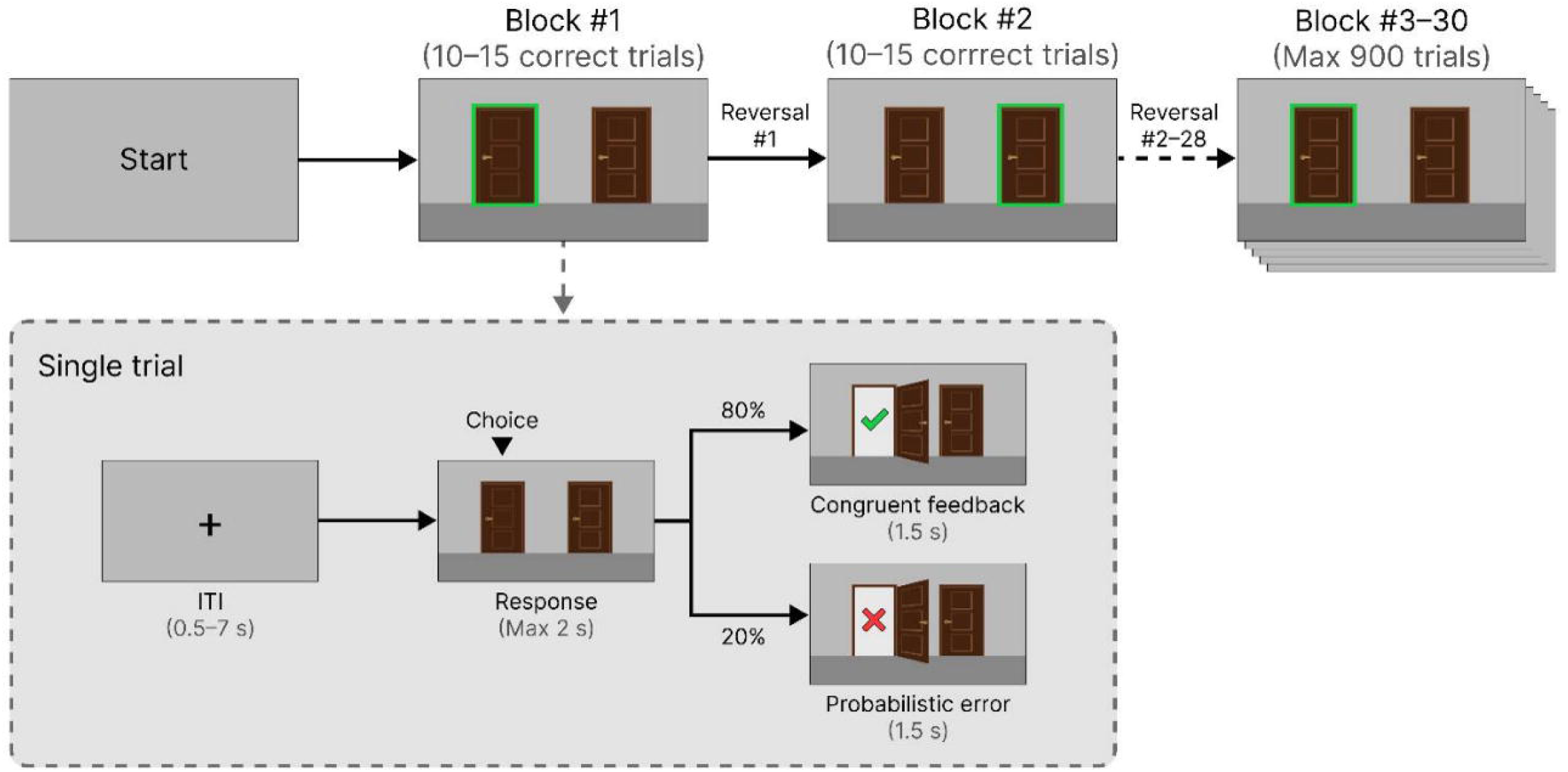
Schematic presentation of the PRL task. ITI = inter-trial interval. Participants are presented with two closed doors and instructed to select the door they believe is correct. Immediate feedback is provided by displaying the selected door open, with either a green checkmark or a red cross behind. Each block follows a contingency rule for a variable number of trials (until 10-15 correct responses), after which the rule reverses without warning such that the previously correct door becomes incorrect and vice versa. Feedback is provided on a probabilistic 80:20 schedule, such that on 20% of randomly selected trials the feedback presented does not reflect the true contingency (probabilistic error). The task consists of 30 blocks (28 reversals) with a maximum of 900 trials (distributed across three 15 min sessions).

Participants are instructed to choose the door they believe is most likely to be correct. Unknown to the participants, feedback is determined according to a probabilistic rule, where one door has 80% chance of giving positive feedback and the other 20% chance. After 10-15 correct responses or a maximum of 30 trials, this probabilistic rule is reversed so that the previous 80% chance becomes 20% chance, and vice versa.

In total, 900 trials are presented over three sessions. The initial correct door is randomized at the start of each session. Extensive pilot testing will be used to inform of any need for adjustment of the number of blocks and proportion of reversals. We will also carry out additional pilot testing in the real scanner. The duration of the task is estimated to be approximately 45-60 min.

Behavioral data will be analyzed focusing on common key metrics for the PRL task, including: 1) proportion of choices towards the currently correct door, 2) reversal adaptation latency, the number of trials needed to shift to the new correct door, and 3) Win-Stay/Lose-Shift. These behavioral metrics will be compared between the two groups and correlated with fMRI-data. Moreover, a number of cognitive models will be fitted to the behavioral data and used to evaluate brain activations in the PRL task.

Existing laboratory facilities at the Department of Psychology, Umeå University are equipped with a mock scanner (realistic model of a real MR scanner with recorded sounds from the real MR environment). Thus, before the fMRI investigation, participants will be primed in this setting.

##### CBF measurement

The integrity of the microcirculation will be assessed by ASL according to recommended guidelines.^34^ During this assessment, the participants will be resting (i.e. no task is involved). Differences in microcirculation between the autistic and neurotypical group will be evaluated.

#### 2. KinArm measurement

In the second step, participants (autistic and neurotypical) will undergo a PRL-paradigm investigating motor flexibility (outside of the MR-scanner). The experiments will be conducted using a robotic manipulandum platform (Kinarm End-Point Robot, BKIN Technologies) and involve a task where a cursor is moved between sequentially appearing targets on a computer screen (Fig 3). Only one target is visible at a time, and the color of this target is related to the probable spatial position of the next target. Given that the overall task goal is to complete as many targets as possible, the color of the visible target can be exploited to predict the location of the next target, and thus to enhance task performance by planning the forthcoming movement in advance.

**Figure 3.**
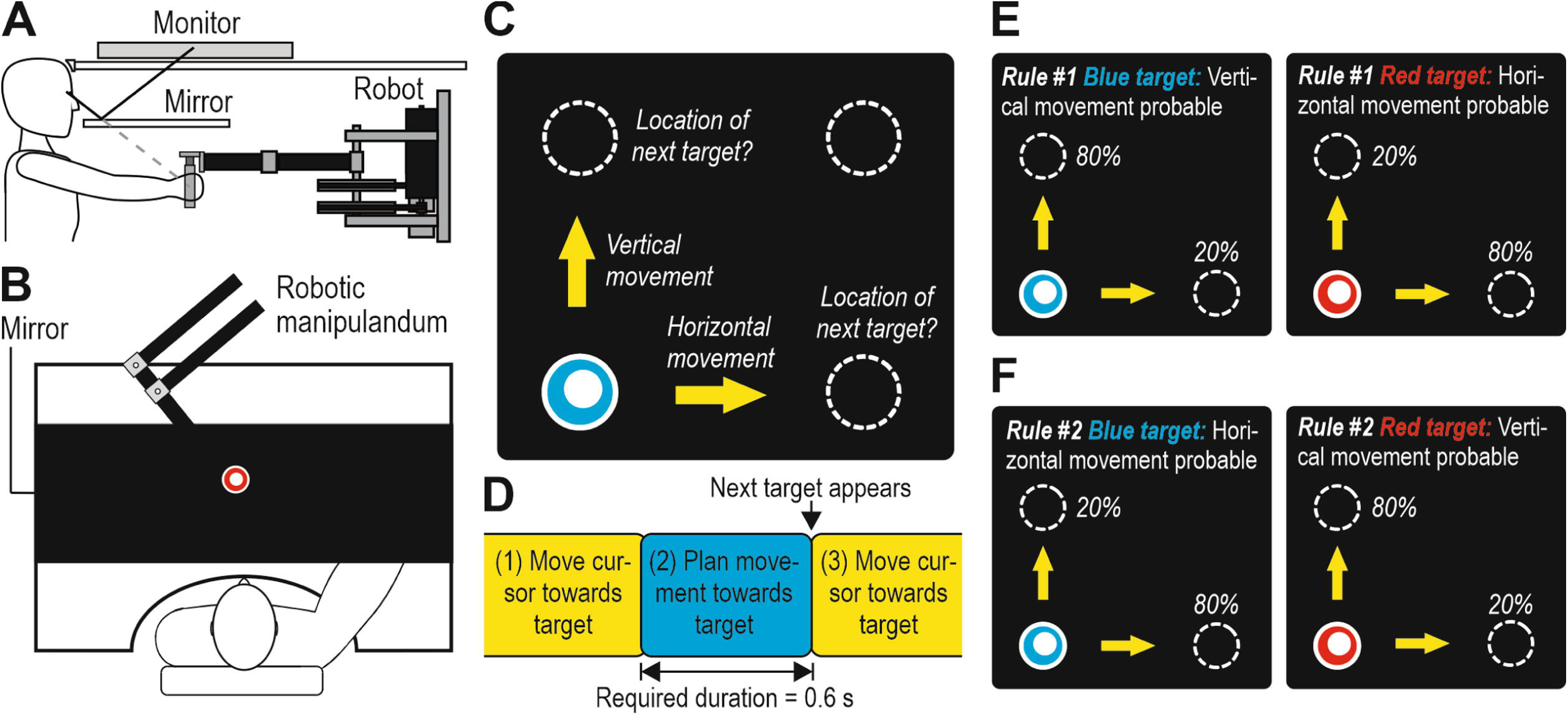
The KinArm End-Point Robot equipment and the motor flexibility experiment. A-B: The cursor and the targets are displayed on a black monitor visible by means of a mirror. The participants can steer the cursor using a handle, but they cannot see their hand or the handle below the mirror. C: The task involves moving a cursor between targets that can be located at four different positions on the screen. However, at any given moment, only one target is visible, and the color of this target is related to the position of the next target. Based on color, it is therefore possible to plan a vertical or horizontal movement towards the next target in advance. D: The task is comprised of sequential action phases of movement execution (when the cursor is moved between targets; represented schematically by yellow boxes, corresponding to yellow arrows in C) and movement planning (when the cursor is held within the visible target; represented by the blue box, corresponding to blue target in C). E-F: According to rule #1, a blue target is followed by a new target above/below the visible target (requiring a vertical movement) with an 80% probability, whereas a red target is followed by a new target left/right of the visible target (requiring a horizontal movement) with an 80% probability. With rule #2, the relationship between color and the probable spatial location of the next target is reversed.

During the experimental session the participants will be chasing targets during sets of 60 trials (about 60 s), separated by 8 s periods of rest. During these 60 trials, the relation between color and the probable spatial position of the next target is unexpectedly reversed every 18-22 trials (Fig 4). In 80% of the trials, the color of the target will correctly indicate the spatial location of the forthcoming next target in accordance with the current rule. In the remaining 20% of the trials the next target will appear at a location in conflict with the current rule, and these trials therefore represent probabilistic errors that must be ignored. Each participant will complete a total of 1400 trials, after which the experiment will end automatically.

**Figure 4.**
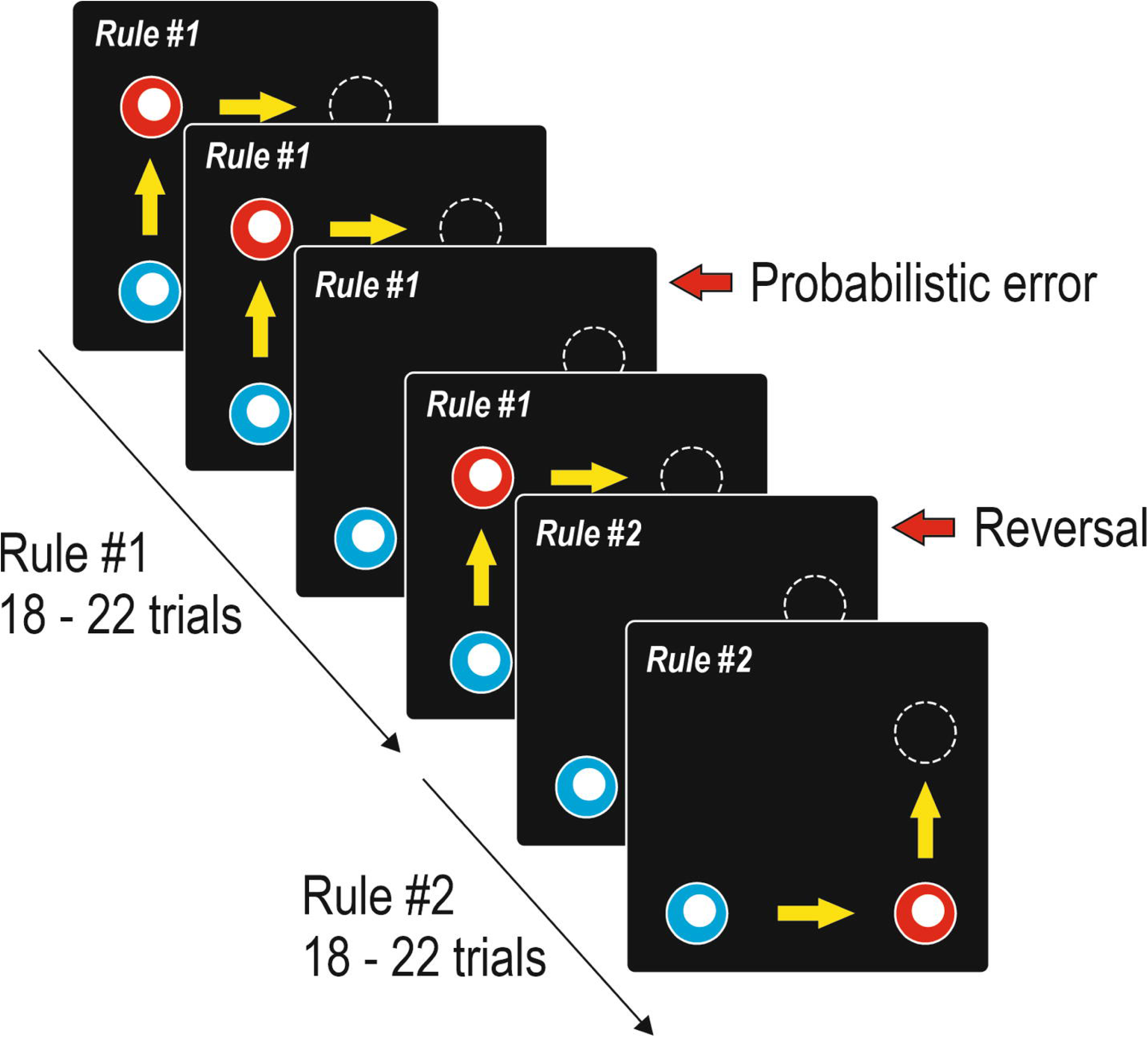
Experimental design. Every 18-22 trials there is a reversal, that is, a rule change between rules #1 and #2. In addition, within each chunk of 18-22 trials, there will also be about 4 probabilistic errors that do not indicate a persistent rule change.

In addition, the same cognitive models as applied for the cognitive PRL task will be fitted to the behavioral data also to evaluate motor behavior.

#### 3. Semi-structured interviews

In the third step, semi-structured interviews with a sample of autistic participants will be conducted. The interview guide encompasses questions falling under four key areas: (1) The ability to be flexible in mind and during problem solving; (2) The ability to coordinate motor behavior and learn movements; (3) Flexible behavior in school and educational settings; and (4) Strategies used to support flexible behavior. Changes over time in all areas are further of interest. The interview will last for approximately 45 min. The collected answers will then be transcribed, coded, and subjected to thematic analysis.

#### Data collection

Full study participation in this project comprises three sessions: a first session for priming and background data collection, one for the fMRI investigation, and one for the KinArm investigation. A fourth session (interview) will be offered to a subset of the autistic participants. Time between sessions may vary although it is not expected to be more than 2 weeks.

##### Priming and background data collection

The first session (Department of Psychology, Umeå University) includes (i) priming in the mock scanner, and (ii) behavioral assessments. Priming in a mock scanner is made to familiarize participants with lying in a MR scanner, and to immediately rule out participation in case of claustrophobia. Behavioral assessments consist of questionnaires and neuropsychological testing. The questionnaires include a demography questionnaire, a Swedish modified version of the Edinburgh Handedness Inventory, a Swedish 50-item version of the Adult Autism Spectrum Quotient (AQ), the Flexibility in Daily Life Scale (FIDL), and the Hospital Anxiety and Depression Scale (HADS). The autism group additionally completes the Repetitive Behavior Scale-Revised (RBS-R). Fluid intelligence is assessed by Matrix Reasoning and Figure Weights from the Wechsler Adult Intelligence Scale, Fourth Edition (WAIS-IV), combined into a fluid reasoning index.^35^ Working memory is assessed by Digit Span Test (WAIS-IV) and an n-back task.^36^ Set shifting is measured using the Trail Making Test Part B (TMT-B) from the Delis-Kaplan Executive Function System (D-KEFS). Fine motor ability is assessed with the Grooved Pegboard Test (Lafayette Instruments). All assessments are performed in a room dedicated for this purpose by, or under supervision of, a licensed psychologist (ED). Outcomes regarding general cognitive and fine motor abilities are of importance to the project due to potential interrelations in autism^37^ and may be included as covariates in some of the analyses.

##### fMRI investigation

For the fMRI investigation, instructions are given to the participant before entering the scanner, and any minor visual impairment is corrected by MR-compatible goggles. A member of the MR staff informs the participant about the MR procedure, and the participant is provided with earplugs and ear protectors to protect from scanner noise. The participant is placed comfortably supine in the scanner. A computer screen, placed behind the scanner bore, is visible through a tilted mirror affixed to the head coil. During the experiment, the screen is used to display the PRL-based paradigm together with written instructions. The participant provides responses to the tasks by pressing one of two designated buttons on a 4-button fMRI response pad (corresponding to the right and left door), held in the right hand. The imaging protocol (same for all participants) includes initial T1-weighted structural imaging (4:11 min), task-based fMRI imaging (3×15 min), T2*-weighted imaging (2:05 min), and the ASL perfusion measurement (5:02 min). The heart rate and respiration of the participant are monitored during scanning. At any time, the participant can communicate with MR staff by using an emergency squeeze ball. The participant is, however, instructed to mainly squeeze this ball if feeling discomfort or in need of stopping the scan for any reason.

##### MRI data acquisition and preprocessing

Image acquisition is made using a 3T GE (General Electric) SIGNA Premiere scanner, with a 48 channel head coil. T2-* weighted images are obtained using a single shot gradient echo planar imaging (EPI) sequence used for BOLD imaging. The following parameters will be used for the imaging sequence: echo time: 30ms, repetition time: 2000ms (410 volumes acquired), flip angle: 80°, field of view: 25×25cm, acquisition matrix: 96×96, reconstruction matrix: 128×128, slice thickness: 3.4mm, slice spacing 0.5, number of slices: 37, slice order: interleaved and in-plane acceleration (ASSET): 2. Ten dummy scans are collected and discarded before the start of the fMRI data collection to allow for equilibrium of the BOLD-signal. T1-weighted images are obtained using a 3D fast spoiled gradient echo sequence (MP-RAGE) in axial orientation. The scan parameters are: echo time: 2.8ms, flip angle: 8°, field of view: 24 x 24cm, acquisition matrix: 240×240, reconstruction matrix: 512×512, slice thickness: 1mm, number of slices: 364, prep time: 1000ms and recovery time: 900ms. In addition, in plane acceleration is achieved by combining compressed sensing with parallel imaging, HyperSense: 1 and ARC: 2.

All images will be corrected for slice timing and realigned to the first image volume of the series. All images will be spatially smoothed through DARTEL.^38^ Segmentation of the individual T1-weighted images will be carried out to generate maps of gray matter (GM), white matter (WM), and cerebrospinal fluid (CSF). Based on these, the DARTEL algorithm will be applied to create a group-specific average template and corresponding individual flow fields.^38^ The resulting DARTEL template and flow field files will then be used to normalize the images to MNI space (2 mm isotropic resolution), after which the images will be smoothed using an 8 mm FWHM Gaussian kernel. Data will also be high-pass filtered to remove low frequency noise. The fMRI data will be analyzed using SPM implemented in Matlab (Mathworks, Inc., USA) and run through an in-house program (DataZ).

##### MRI data analysis

For fMRI data analysis, whole-brain and region-of-interest analyses (fronto-striatal regions) will be used. The following contrasts will be of interest: (1) first reversal error after previous congruent feedback contrasted against correct responses with congruent feedback (2) final reversal errors contrasted against correct responses with congruent feedback (3) other preceding reversal errors contrasted against correct responses with congruent feedback, (4) probabilistic errors contrasted against correct responses with congruent feedback.

At the individual level, a general linear model of all events of interest will be set up from onset to offset. In addition, the six movement parameters and ITI (modeled as onset to offset of the event) will be used as regressors of no interest in the model. At the second level, group analysis will be performed separately for each group, and differences between the autism and neurotypical groups will be performed using univariate (*t*-tests, ANOVAs, brain-behavior correlations) and multivariate (representational similarity analysis, multivoxel pattern analysis) methods. Model-based fMRI will additionally be used to evaluate how predictions in performance relate to brain activation and related to differences between the two groups.^33^

##### KinArm investigation

For the KinArm investigation (Department of Medical and Translational Biology, Umeå University), the participants will be explicitly instructed to complete as many targets as possible during each period of target chasing. This is to ensure that they will attempt to exploit the rule between target color and the probable location of the forthcoming target to plan the forthcoming movement in advance and thus enhance performance. To motivate the participants, during the rest periods, the screen will show the number of completed targets during the previous target-chasing period, and the “high score” of targets completed during any previous period. While performing the task, the participants will sit on a height-adjustable chair in front of the robotic manipulandum platform with their forehead against a support. They will control the cursor with their preferred (dominant) hand using a handle attached to the robot. The cursor is centered on the handle, but the participants cannot see the handle or their hand beneath the mirror.

In the KinArm experiments, spatiotemporal kinematic data about the cursor movements will be sampled at 1000 Hz. Of particular interest is the duration of movement executions between targets (see Fig 3C-D), because this measure will indicate how the participants use the color of the targets (the sensorimotor rules) to support the planning of forthcoming movements. Thus, the data analysis will focus on group differences in how sensorimotor rules are established and updated in case of rule changes and probabilistic errors. This will enable a sensitive comparison of the flexibility in motor performance between the autistic and neurotypical participants.

##### Interview study

The interview session, offered to autistic participants, consists of an individual interview conducted digitally via a secure University-licensed version of Microsoft Teams. The interviews will be carried out by an experienced qualitative researcher (MEN). Upon agreement on a date and time of their own choice, participants will receive a personalized Microsoft Teams invitation containing a unique meeting link. The meeting will be accessible only to the researcher and the respective participants. The interview will be recorded using the built-in recording function in Microsoft Teams, and transcription will be generated directly through the secure Teams transcription function. Both recordings and transcripts will be stored within the University’s password-protected and access-restricted Teams environment. As participation in the interview study does not require participation in other project investigations, some autistic participants may be recruited exclusively for this qualitative component.

The interview study will explore perceived changes over time among the participants, regarding cognitive flexibility and problem-solving, fine and gross motor skills, in educational contexts, and during leisure time. It will also examine strategies used to support flexible behavior. Outcomes will be reported as themes and patterns identified through thematic analysis of semi-structured interview data.

## Data Availability

As required by GDPR, access to de-identified data files from the present study will be available upon request pending University legal counselling

## ETHICS AND DISSEMINATION

The project has been approved by the Swedish Ethical Review Authority (ref:2025-07939-01) and will be conducted in accordance with the Declaration of Helsinki. All investigations described are well-established, non-invasive and safe to partake in. Participants are also explicitly informed about the possibility to terminate participation at any time without fear of repercussion. In addition, the project includes classification, risk- and vulnerability analysis based on best practice according to the Swedish Civil Contingencies Agency, and a plan for data storage/liquidation. In accordance with the European Union General Data Protection Regulation (GDPR) and national guidelines for the storing of personal data, an exclusive secure file area governed by the Umeå University IT department has been set up for the project. Presently, no datasets have been generated or analyzed. However, when practically possible, data will be de-identified in accordance with GDPR and Data Protection Act 2018. All data will be pseudonymized by a specific study code applied to each participant. Brain imaging data will be stored in a secure computer database kept at the UFBI. Additional background and behavioral data collected outside the scanner will be kept securely in a fire-proof safety cabinet in a locked laboratory at the Department of Psychology, Umeå University. Only those directly involved in the project will have access to data. Outcomes from this project will be shared with the scientific community in the shape of publications in peer-reviewed international scientific journals (open access), disseminated at both national and international conferences, and presentations to any interested audience/stakeholders. As required by GDPR, however, access to de-identified data files will only be available upon request, pending University legal counselling.

## AUTHORS’ CONTRIBUTION

**Conceptualization:** Erik Domellöf, Andreas Johansson, Sara Stillesjö, Linnea Karlsson Wirebring, Carola Wiklund-Hörnqvist, Anna-Maria Johansson, Thomas Rudolfsson, Anders Wåhlin, Daniel Säfström

**Data curation:** Erik Domellöf, Andreas Johansson, Sara Stillesjö, Linnea Karlsson Wirebring, Carola Wiklund-Hörnqvist, Anna-Maria Johansson, Thomas Rudolfsson, Anders Wåhlin, Gustaf Wadenholt, Malin Ekesryd Nordström, Daniel Säfström

**Formal analysis:** Erik Domellöf, Andreas Johansson, Sara Stillesjö, Linnea Karlsson Wirebring, Carola Wiklund-Hörnqvist, Anna-Maria Johansson, Thomas Rudolfsson, Anders Wåhlin, Gustaf Wadenholt, Malin Ekesryd Nordström, Daniel Säfström

**Funding acquisition:** Erik Domellöf, Carola Wiklund-Hörnqvist, Anna-Maria Johansson

**Investigation:** Erik Domellöf, Andreas Johansson, Sara Stillesjö, Linnea Karlsson Wirebring, Carola Wiklund-Hörnqvist, Anna-Maria Johansson, Thomas Rudolfsson, Anders Wåhlin, Gustaf Wadenholt, Malin Ekesryd Nordström, Daniel Säfström

**Methodology:** Erik Domellöf, Andreas Johansson, Sara Stillesjö, Linnea Karlsson Wirebring, Carola Wiklund-Hörnqvist, Anna-Maria Johansson, Thomas Rudolfsson, Anders Wåhlin, Gustaf Wadenholt, Malin Ekesryd Nordström, Daniel Säfström

**Project administration:** Erik Domellöf, Andreas Johansson, Sara Stillesjö, Linnea Karlsson Wirebring, Carola Wiklund-Hörnqvist, Anna-Maria Johansson, Thomas Rudolfsson, Anders Wåhlin, Gustaf Wadenholt, Malin Ekesryd Nordström, Daniel Säfström

**Software:** Andreas Johansson, Sara Stillesjö, Linnea Karlsson Wirebring, Gustaf Wadenholt, Daniel Säfström

**Supervision:** Erik Domellöf, Sara Stillesjö, Anders Wåhlin, Daniel Säfström

Writing - original draft: Erik Domellöf

**Writing – review and editing:** Erik Domellöf, Andreas Johansson, Sara Stillesjö, Linnea Karlsson Wirebring, Carola Wiklund-Hörnqvist, Anna-Maria Johansson, Thomas Rudolfsson, Anders Wåhlin, Gustaf Wadenholt, Malin Ekesryd Nordström, Daniel Säfström

